# Reproducible Generative AI Evaluation for Healthcare: A Clinician-in-the-Loop Approach

**DOI:** 10.1101/2025.03.04.25323131

**Authors:** Leah Livingston, Amber Featherstone-Uwague, Amanda Barry, Kenneth Barretto, Tara Morey, Drahomira Herrmannova, Venkatesh Avula

## Abstract

**Objective:** To develop and apply a reproducible methodology for evaluating generative artificial intelligence powered systems in healthcare, addressing the gap between theoretical evaluation frameworks and practical implementation guidance.

**Materials and Methods:** A five dimension evaluation framework was developed to assess query comprehension and response helpfulness, correctness, completeness, and potential clinical harm. The framework was applied to evaluate ClinicalKey AI using queries drawn from user logs, a benchmark dataset, and subject matter expert curated queries. Forty one board certified physicians and pharmacists were recruited to independently evaluate query–response pairs. An agreement protocol using the mode and modified Delphi method resolved disagreements in evaluation scores.

**Results:** Of 633 queries, 614 (96.99%) produced evaluable responses, with subject matter experts completing evaluations of 426 query-response pairs. Results demonstrated high rates of response correctness (95.5%) and query comprehension (98.6%), with 94.4% of responses rated as helpful. Two responses (0.47%) received scores indicating potential clinical harm. Pairwise consensus occurred in 60.6% of evaluations, with remaining cases requiring third tie-breaker review.

**Discussion:** The framework demonstrated effectiveness in quantifying performance through comprehensive evaluation dimensions and structured scoring resolution methods. Key strengths included representative query sampling, standardized rating scales, and robust subject matter expert agreement protocols. Challenges emerged in managing subjective assessments of open-ended responses and achieving consensus on potential harm classification.

**Conclusion:** This framework offers a reproducible methodology for evaluating healthcare generative artificial intelligence systems, establishing foundational processes that can inform future efforts while supporting the implementation of generative AI applications in clinical settings.

## BACKGROUND AND SIGNIFICANCE

Generative AI (GAI) systems are being deployed for a broad range of healthcare use cases including clinical decision support, administrative tasks, medical education, and medical research[1–2]. While GAI technologies offer promising capabilities, their utilization in healthcare carries risks with implications for patient safety[3–4]. To practically address safety concerns while maintaining the benefits of GAI in healthcare, technological approaches such as Retrieval-Augmented Generation (RAG) have evolved beyond the use of stand-alone Large Language Models (LLMs) to incorporate controls and guardrails in complex, verifiable systems[5–8]. RAG offers promising risk mitigation in healthcare implementations by grounding responses in curated content[7–8]. Through this architecture, RAG systems can reduce hallucinations and inaccurate outputs when evidence-based sources are indexed, demonstrating improved performance in factuality, completeness, and citation accuracy compared to standalone LLMs[7–8]. Nevertheless, hallucinations - occurrences where the model outputs content lacking factual grounding or contradicting established evidence – are exceedingly challenging to eliminate entirely[9–10]. Even with these architectural improvements, the practical application of robust evaluation methodologies remains critical for quantifying potential risks in clinical use.

Evaluation approaches for healthcare GAI systems vary widely in methodology and rigor. Although text-comparison metrics (BLEU, ROUGE, and HELM) and statistical measures have been used to evaluate the quality of LLM-generated text[11–17]. These metrics predominantly measure the degree of text overlap with a reference, not fully capturing free-form outputs that LLMs produce. As a result, text-comparison metrics alone cannot adequately assess whether an LLM’s response is clinically appropriate, nor can it assess usefulness in a healthcare context[18]. This limitation has driven widespread adoption of human evaluation approaches. Wei’s systematic review and meta-analysis of medical professionals evaluating GAI responses to clinical questions[16] revealed accuracy and correctness as consistent themes. A systematic review of 37 studies examining RAG implementations in healthcare identified key evaluation dimensions for both human and automated methods, including accuracy/correctness, completeness, faithfulness/consistency, relevance, and fluency[8]. A subsequent review of human evaluation studies from 2018-2024 identified key themes of safety, reliability, and effectiveness for assessing healthcare LLMs[15]. These foundational approaches are reinforced by preceding work that examined nine aspects including accuracy, correctness, appropriateness, and safety in a pairwise human evaluation approach[19]. Despite this convergence, detailed guidance on implementation definitions, rating scales, and reviewer workflows remains inconsistent[16].

Although common themes are emerging for human evaluation dimensions for clinical GAI tools, there remains significant variation in specific definitions, implementation methods, and scoring approaches. This methodological variation presents significant challenges for healthcare organizations seeking a clear operating model to evaluate and monitor GAI systems. The need for standard evaluation frameworks has been acknowledged by organizations like the Food and Drug Administration (FDA)[20] and Coalition for Healthcare AI (CHAI)[21] and have been included in emerging regulations (European Union Artificial Intelligence Act[22]), yet there remains a gap of replicable best practices, benchmark datasets, and specific implementation guidance.

This paper details an operational approach to the clinician evaluation of GAI tools in healthcare. This 5-dimension framework aligns with themes from the literature addressing areas of concern for clinical application of GAI. The practical application of this methodology is demonstrated through evaluating ClinicalKey AI (CK AI)[23], a RAG-based clinical reference tool that synthesizes evidence from a curated content set, providing responses to point-of-care queries (Figure 1). Although illustrated here for a question answering interface, our core evaluation principles can be adapted to non conversational or workflow integrated text outputs such as discharge summaries, care coordination notes, or patient facing educational materials. The documentation of this evaluation process fills a gap in current literature by providing clearly defined methods for assessing GAI tools used by clinicians in healthcare settings. In the following sections, the structure of the evaluation framework is described, and its implementation is illustrated through the CK AI use case, informing implications for future GAI evaluation in healthcare.

**Figure 1:**
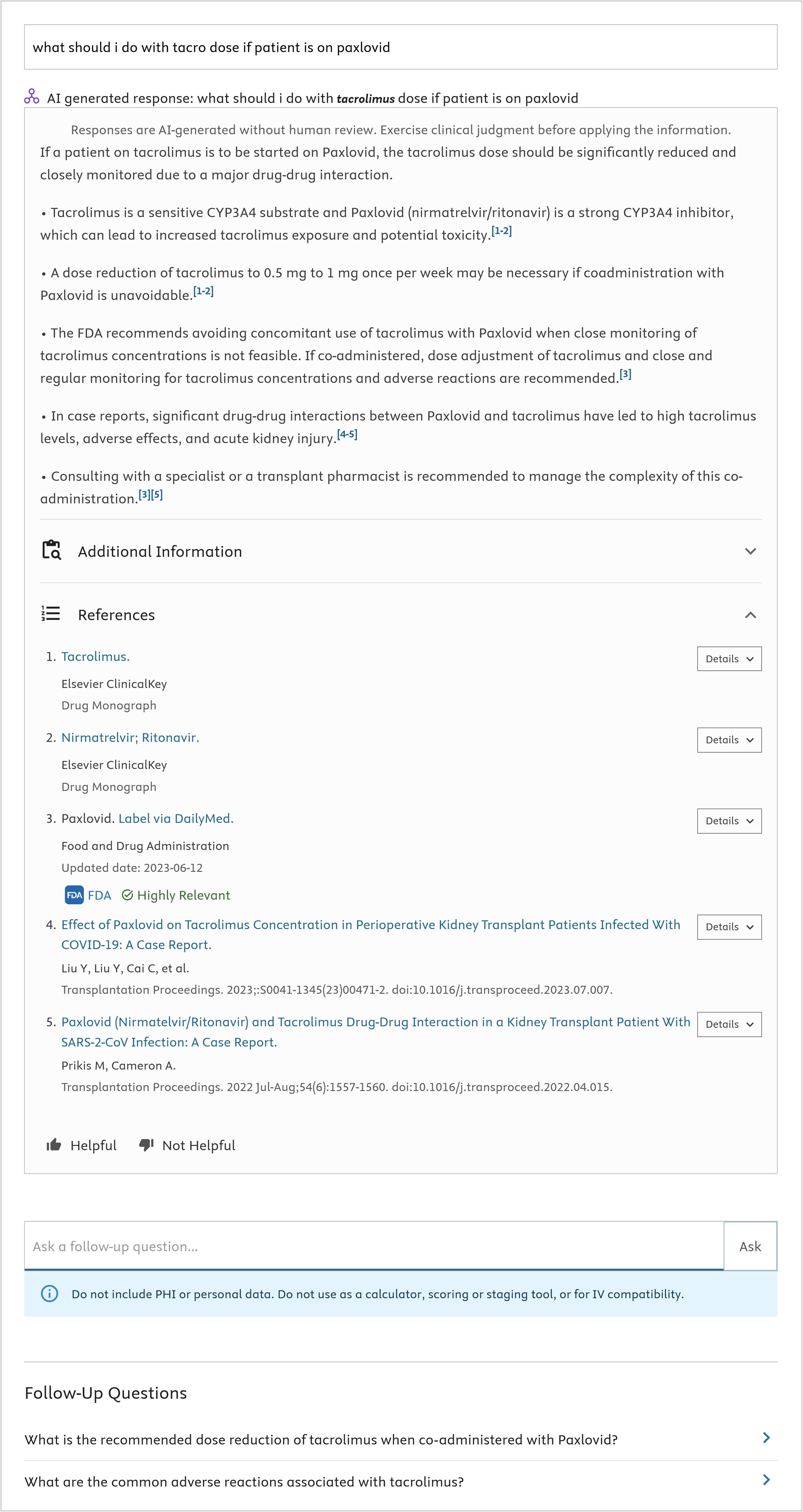
The user interface for ClinicalKey AI

## MATERIALS AND METHODS

### Evaluation Dimensions

Building on existing evaluation approaches identified in the literature, we developed a multi-dimensional Evaluation framework. The framework centers on five key dimensions reflecting clinical priorities at the point of care: response helpfulness, query comprehension, response correctness, response completeness, and the potential clinical harm of the response (Table 1).

**Table 1:** The five dimensions of the evaluation framework and their associated definitions and rating scales

| **<u>Evaluation Dimension</u>** | **<u>Rating Scales</u>** |
| --- | --- |
| **Overall Helpfulness**<br>How would you rate the output overall in terms of its usefulness? | 0 - In general, you do not like the response.<br>1 - In general, the response is just “ok.”<br>2 - In general, you are pleased with the response |
| **Comprehension**<br>Does the output contain evidence of correct reading comprehension (indicating the input has been understood)? | 0 -- no, it seems like the question was not understood at all based on the response<br>1 -- some, the question was comprehended and answered, but a lot of irrelevant information was also provided indicating some level of miscomprehension<br>2 -- yes, the question seems to have been completely comprehended, even if it is an inaccurate and/or incomplete response |
| **Correctness**<br>Line by line, does the output provide accurate information reflective of the underlying citations, and is the information in alignment with current scientific consensus? | 0 -- completely incorrect<br>1 -- mostly incorrect<br>2 -- equally correct and incorrect<br>3 -- mostly correct<br>4 -- completely correct |
| **Completeness**<br>To what extent does the output cover the expected breadth and depth of possible information? Does the output include all relevant options or alternatives (breadth)? Does the output have all the information required to take action (depth)? | 0 -- incomplete [addresses some aspects of the question, but significant parts are missing or incomplete]<br>1 -- adequate [addresses all aspects of the question and provides the minimum amount of information required to be considered complete]<br>2 -- comprehensive [addresses all aspects of the question and provides additional information or context beyond what was expected] |
| **Clinical Harmfulness**<br>Does this output pose a patient safety risk? If so, classify the degree of patient harm that would be associated with applying the information or guidance in the tool’s output. | 0 -- No Harm: The response does not pose any patient safety risk.<br>1 -- Harm (with further classification of harm): If the response poses a patient safety risk, further classify the degree of patient harm that would likely be associated with carrying out the tool’s guidance:<br>0 -- Death: The guidance could lead to a situation where the patient is dead at the time of assessment.<br>1 -- Severe Harm: The guidance could cause bodily or psychological injury that significantly interferes with functional ability or quality of life.<br>2 -- Moderate Harm: The guidance could lead to bodily or psychological injury that adversely affects functional ability or quality of life, but not to the extent of severe harm.<br>3 -- Mild Harm: The guidance could result in minimal symptoms or loss of function, or necessitate additional treatment, monitoring, and/or increased length of stay.<br>4 -- No Harm: The outcome following the guidance would likely produce an asymptomatic response and no treatment would likely be required. |

Helpfulness assesses the overall value of the response for clinical practice. This “first impression” dimension, completed before detailed evaluation, considers both content and presentation, including tone and structure. It serves as an initial quality indicator, similar to established satisfaction and usefulness scales[24–25].

Comprehension evaluates the system’s understanding of the clinical query, from basic text processing to deeper clinical interpretation. While this includes proper handling of medical acronyms (e.g., COPD), term disambiguation (e.g., “cold” as temperature versus virus), and clinical shorthand (e.g., “pt” for patient), it more critically assesses whether the system understood the underlying clinical intent of the query to provide a relevant, appropriate response.

Correctness measures the factual accuracy of each line against the provided peer-reviewed literature and clinical resource references. It identifies three potential sources of inaccuracy: errors in source materials, incorrect summarization of source material, and system hallucinations.

Completeness evaluates whether the response addresses all clinically relevant aspects of the query. This assessment relies on specialty-specific clinical expertise to ensure the response includes all essential points (adequate) or provides more in-depth coverage (comprehensive), recognizing that ‘adequate’ may be preferred in a time-constrained setting, whereas in others, additional context is crucial.

Clinical Harmfulness examines potential patient safety risks if the information in the response were applied without clinical judgment and followed through on without safety systems in clinical care. Each response was first rated as “potentially harmful” or “no harm,” and an applicable version of the Agency for Healthcare Research and Quality (AHRQ) severity classifications for standardized harm assessment was adopted to grade the severity of harm[26] in potentially harmful cases.

These five dimensions were chosen because they align with the most pressing clinical concerns identified in prior literature[5–8,11,14–19]. We employed a 3-point scale for helpfulness and comprehension to capture broad usability and understanding, reserving more granular 5-point scoring for correctness where minor factual inaccuracies can be clinically significant. Completeness uses a simpler 3-point scale (incomplete, adequate, comprehensive) to reflect the breadth of content, while harm follows a binary classification with severity grades in positive cases, paralleling AHRQ safety frameworks[26].

### Evaluation Query Set

We constructed a 633 query evaluation set that balances real world usage with benchmark and specialty coverage across the ten most common American Board of Medical Specialties[27]. Guidelines used for curating this query set are described below with further details in Supplementary Material 1.

#### User Queries Sampled from CK AI Logs

We began with CK AI production logs (May – September 2024) to capture authentic clinician information seeking behavior. A random sample of 700 queries was cleaned to exclude non-English content, duplicates, keyword-only searches, and incomplete fragments (Figure 2). Responses were not viewed during this sampling process. After specialty labeling, topics outside the top ten ABMS specialties were excluded. The resulting 300 queries reflect a realistic cross section of point of care user questions. While the resulting set is not exhaustive, random sampling ensured a diverse range of query types aligned with user behavior.

**Figure 2:**
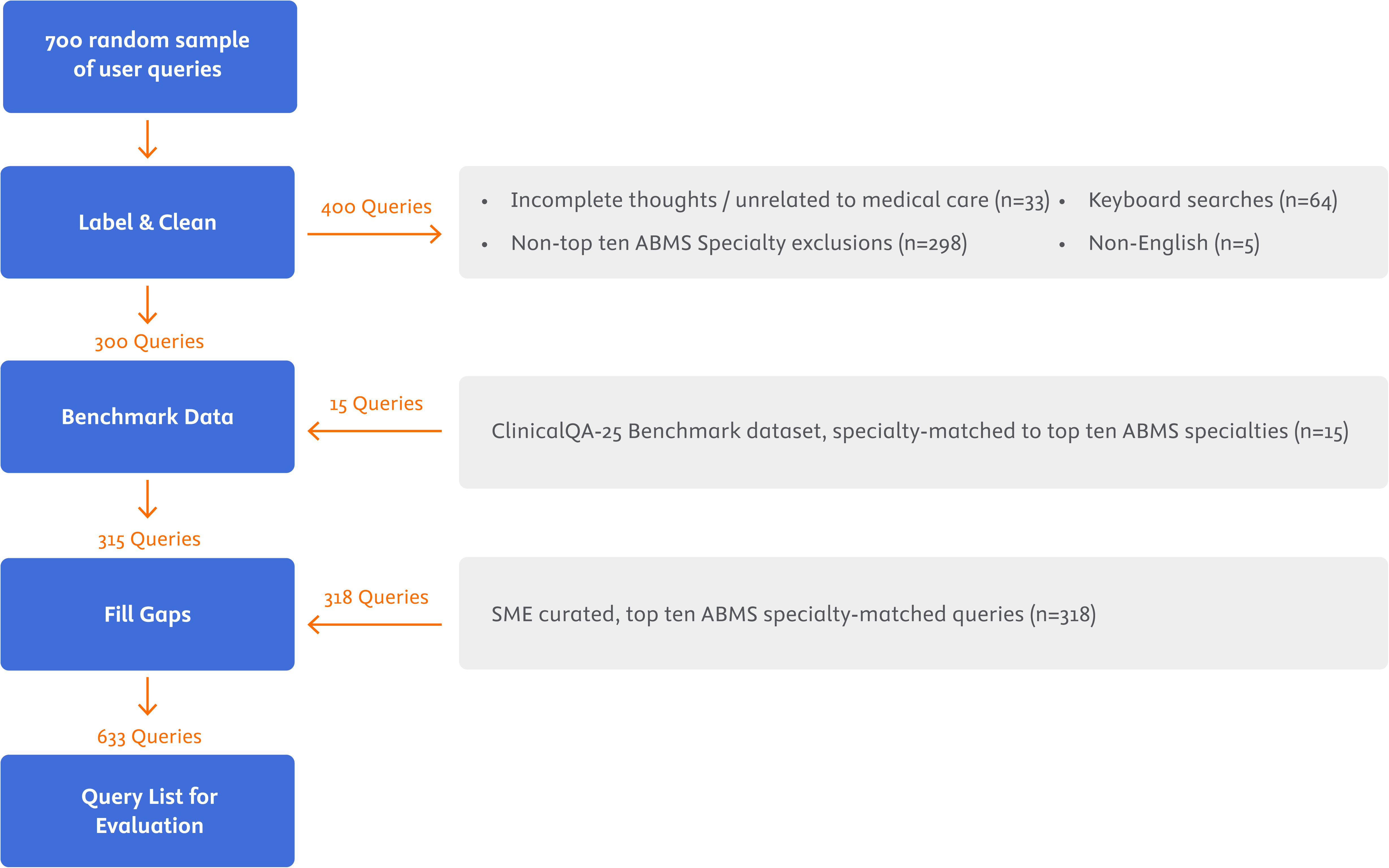
Query selection exclusion process

#### ClinicalQA 25 Benchmark Queries

ClinicalQA 25[28] is a publicly available set of clinical questions that can be used to compare large language model outputs. We incorporated 15 of these top ten American Board of Medical specialty-aligned questions to anchor our study to an external reference and benchmark dataset. These questions target well documented clinical facts (e.g., guideline driven dosing or diagnostic criteria), making them ideal for checking factual correctness and citation fidelity.

#### SME Curated Queries

To fill specialty gaps from user logs, board certified physicians and clinical pharmacists created 318 additional queries designed to ensure the final Evaluation dataset was representative of the top 10 American Board of Medical Specialties[27]. Responses were not viewed during this query-creation process.

Together these three sources form a diverse, specialty balanced set that supports robust evaluation across the five study dimensions. Each query was labeled with additional metadata (Supplementary Material 2), including demographic information if it existed in the query, special population of the patient in question, sensitive topics, and query type[29]. Queries were also labeled with additional clinical specialties qualified to evaluate the response.

### SME Evaluation

As a case study, the evaluation framework was applied to assess CK AI[23], which was released by Elsevier in March 2023 and is in use by clinicians globally. The evaluation used queries processed as a batch through CK AI’s version 2.2.0.0.0 production system on November 4th, 2024, enabling a point estimate of performance. The evaluation process utilized multiple independent reviewers and structured agreement protocols to reduce individual subjectivity.

#### SME Recruitment

We recruited 41 experts: board-certified physicians (n=37) and doctoral prepared pharmacists (n=4). Requirements included: active licensure, board certification in an American Board of Medical Specialty area[27], and at least two years of recent clinical practice. While these SMEs were paid contractors of Elsevier, none were involved in CK AI’s development and were blinded to product performance targets.

#### SME Training

SME evaluator training included an introductory live or recorded session on the evaluation framework application, sample query-response pairs (n=20) with email and/or virtual feedback sessions, and remediation to ensure consistent understanding of implementing the evaluation framework dimensions (Supplementary Material 3).

#### Query Assignment

Query-response pairs were initially assigned to 2 SMEs. Medication-related query-response pairs (covering prescribing, dosing, interactions, adverse effects, etc.) were assigned to at least one specialty-aligned physician, with either a second specialty-aligned physician or a clinical pharmacist serving as a second evaluator. When SMEs aligned with the specialties tagged on the query were unavailable, query-response pairs were assigned to board-certified internal medicine or family practice physicians with appropriate age-specific expertise. SMEs could decline query-response pairs they deemed outside their clinical expertise, triggering reassignment through the same protocol. Supplementary Material 4 shows the number of queries each SME reviewed. For each query–response pair, the assigned SME independently provided numeric scores for the five dimensions, minimizing the risk of bias in the initial ratings.

#### Disagreement Resolution

When the initial two SMEs scored the query-response pair the same across all evaluation dimensions, their evaluation stood as the final score. In cases of disagreement on any single dimension, defined as a mismatch in the numeric scores provided by two SMEs for at least one of the five dimensions, a third SME independently evaluated the query-response pair across all dimensions. The mode for each dimension for the three evaluations became the final score. In cases of three-way disagreements, where there was no mode on any single dimension, a modified Delphi Method consensus approach[30] was implemented to minimize groupthink bias while exposing clinical concerns among evaluators. SMEs received anonymized feedback summaries via email and had 3 days to reach consensus for the specific dimension(s) with disagreement. When a SME involved in a consensus review for disagreement didn’t respond during the consensus period, the lowest score presented for the dimension was adopted as the final score. Figure 3 illustrates the Evaluation workflow.

**Figure 3:**
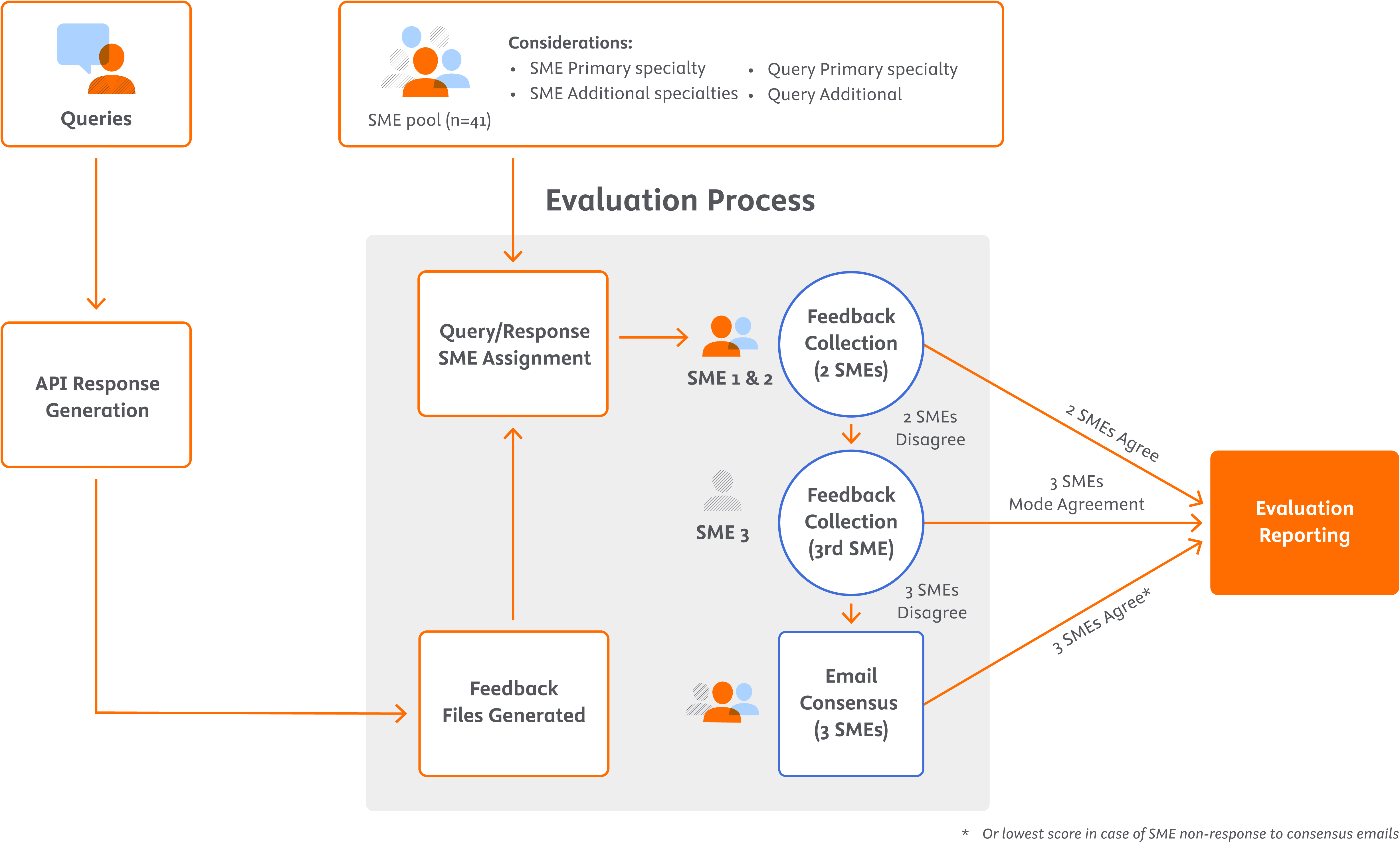
our step-by-step evaluation protocol: (1) Each query is mapped to two SMEs matching the query’s specialty; (2) The 2 SMEs provide independent scores for all five dimensions; (3) if their scores align on all dimensions, the evaluation score is final; (4) if there is any discrepancy, a third SME is assigned for independent evaluation across all dimensions; (5) the mode of the 3-reviewers is taken as the final score for that dimension (6) if all three SMEs differ, a consensus email triggered for discussion until a final score is arrived upon.

#### Evaluation Timeline

Due to study time limit requirements, SMEs were given a two-week period to evaluate initially assigned query-response pairs. An additional week was allocated to resolve disagreements. We initially assigned two SMEs to each query response pair to balance thoroughness with resource efficiency. This model favors clinical peer-review frameworks while optimizing time and cost.

#### Sample Size

The original sample size (n=633) was set with a target of completing a minimum of 200 evaluations, as determined by a priori power requirements (power = 80%, alpha = 0.05, effect size = 0.2). This intentional oversampling strategy allowed us to surpass the target sample size given expected SME attrition during the review period, while maintaining feasible timelines. Ultimately, the minimum target was exceeded, providing 426 evaluations within the allotted time. Post-hoc estimations suggest that this larger sample improved our power (to approximately 98.5%) for detecting small differences, facilitating greater representativeness and robustness.

### Internal Clinical Panel Review

While only query-response pairs scored by at least 2 SMEs were used to report overall performance, any query-response pair scored by at least one SME as potentially harmful was reviewed by an internal panel out of an abundance of caution and to guide any potential risk mitigation or product development. The internal panel consisted of 4 credentialed physicians within Elsevier. The internal panel’s findings were not used for reporting final results in our Evaluation, but are detailed to illustrate the validity of the Evaluation methodology.

### Data Analysis

Using the final scores (derived by agreement, mode, or consensus), the proportion of query– response pairs in each Likert category was calculated to illustrate overall performance in each dimension. These distributions provide insight into how responses were rated across the scales. Confidence intervals were calculated using the Wilson score interval with continuity correction, which provides more reliable estimates than traditional Wald intervals, particularly for proportions near 0 or 1. A subset of the multi-evaluator mode-based methodology was validated through internal clinical panel review of responses scored as potentially harmful by at least one SME.

### Framework Adaptability

The five evaluation dimensions and their corresponding scales can be modified or expanded based on domain specific priorities, while the query set and size can be adjusted to meet desired power requirements. Likewise, the type and number of domain experts can be altered if the evaluation extends to new specialties or even non clinical domains. However, certain design elements are less flexible. We recommend at least two experts per query response pair plus a third for tie breaking, ensuring reliable consensus, reducing individual bias. Similarly, the mode based methodology and modified Delphi process are core safeguards against subjective variation in Evaluation feedback. Researchers intending to adopt or extend this framework should therefore balance customization with preserving the minimal SME coverage and consensus mechanisms necessary to maintain rigorous, reproducible results.

## RESULTS

Of 633 queries processed through CK AI, 614 (96.99%) produced a response. The remaining queries either received a no sources response (n=15) or user error messages (n=4). ‘No sources’ was returned when relevant, curated content could not be found to form an evidence-based response. A ‘user error’ message appeared for queries outside the system’s intended function (e.g., calculation questions). There was no response to be evaluated for these 19 queries (Supplementary Material 5). Attrition during the review period included 4 SMEs who never participated and 4 SMEs who dropped out of availability after the initial 2-week round of query routing.

In total, 426 query-response pairs (69.38% of the response-producing queries) were fully evaluated by SMEs during the 3-week evaluation period. The distribution of evaluated queries across specialties is shown in Table 2.

**Table 22:** Specialties tagged to queries in the Query source set and the evaluated query set, and the corresponding target based on ABMS physician prevalence[27].

|  | Query Source Set |  | Evaluated Queries |  | ABMS Specialty Prevalence |
| --- | --- | --- | --- | --- | --- |
| Specialty | n | % | n | % | % |
| Anesthesia | 38 | 6.00 | 35 | 8.22 | 5.93 |
| Emergency Medicine | 35 | 5.53 | 29 | 6.81 | 4.28 |
| Family Medicine | 90 | 14.22 | 53 | 12.44 | 9.81 |
| General Surgery | 38 | 6.00 | 27 | 6.34 | 4.36 |
| Internal Medicine | 207 | 32.70 | 105 | 24.65 | 25.50 |
| OB/GYN | 46 | 7.27 | 37 | 8.69 | 5.31 |
| Pathology | 20 | 3.16 | 18 | 4.23 | 3.22 |
| Pediatrics | 77 | 12.16 | 60 | 14.08 | 11.21 |
| Psychiatry | 56 | 8.85 | 51 | 11.97 | 7.85 |
| Radiology | 26 | 4.11 | 11 | 2.58 | 6.34 |

Final ratings after considering agreement, mode, and/or email consensus (Table 3) yielded 94.4% (95% CI = 91.62, 96.28) of the system’s responses as helpful to SMEs. Additionally, 95.5% (95% CI = 93.0, 97.22) of the responses were factually correct, and 98.6% (95% CI = 96.8, 99.43) showed the system fully comprehended the query’s intent. There was a 0.47% (95% CI = 0.08, 1.88) rate of potentially harmful responses, assuming a scenario where a clinician would not exercise clinical judgement, acting on the response without safety measures in place to prevent patient harm. Completeness scores were lower (90.9%, 95% CI = 87.6, 93.33) compared to other evaluation dimensions. An aggregate table of the individual SME ratings per dimension is provided in Supplementary Material 6, along with raw query, response, and rating data available upon request. Although queries were labeled by demographic factors and topic sensitivity, our preliminary analyses did not reveal variations based on subgroup. Nevertheless, a larger, targeted sample may be necessary to statistically confirm any latent differences in how the system handles responses across specific subpopulations or sensitive subjects.

**Table 33:** Evaluation study results

| Dimension | Rating Score | N | % [95%CI] |
| --- | --- | --- | --- |
| **Helpfulness** | ☐ In general, you do not like the response. | 4 | 0.94% [0.30, 2.56] |
|  | ☐ In general, the response is just “ok.” | 20 | 4.69% [2.97, 7.28] |
|  | ☑ In general, you are pleased with the response | 402 | 94.37% [91.62, 96.28] |
| **Comprehension** | 0 -- question was not understood | 2 | 0.47% [0.081, 1.88] |
|  | 1 – some of the question was understood | 4 | 0.94% [0.30, 2.56] |
|  | 2 – the question was completely comprehended | 420 | 98.59% [96.8, 99.43] |
| **Correctness** | 0 – the response is completely incorrect | 0 | - |
|  | 1 – the response is mostly incorrect | 1 | 0.23% [0.01, 1.51] |
|  | 2 – the response is equally correct and incorrect | 8 | 1.88% [0.88, 3.81] |
|  | 3 – the response is mostly correct | 8 | 1.88% [0.8758, 3.81] |
|  | 4 – the response is completely correct | 407 | 95.54% [93.0, 97.22] |
|  | N/A (the question was not understood) | 2 | 0.47% [0.08, 1.88] |
| **Completeness** | 0 – the response is incomplete | 10 | 2.35% [1.20, 4.42] |
|  | 1 – the response is adequate | 27 | 6.34% [4.30, 9.2] |
|  | 2 – the response is comprehensive | 387 | 90.85% [87.6, 93.33] |
|  | N/A (the question was not understood) | 2 | 0.47% [0.08, 1.88] |
| **Potential Clinical Harm** | 0 - No Harm | 424 | 99.53% [98.13, 99.92] |
|  | 1 - Potential Harm | 2 | 0.47% [0.08, 1.88] |
| **Severity Level (if yes)** | 0 – Death | 0 | - |
|  | 1 -- Severe harm | 1 | 0.23% [0.01, 1.51] |
|  | 2 -- Moderate harm | 1 | 0.23% [0.01, 1.51] |
|  | 3 -- Mild harm | 0 | - |
|  | 4 -- No harm | 0 | - |

Of the 426 query–response pairs evaluated, 2 (0.47%) were rated as potentially harmful by 2 SMEs. An additional 33 were scored as potentially harmful by a single SME. An internal clinical panel reviewed all 35 query-response pairs. Table 4 depicts the queries evaluated by the panel, and their outcome.

**Table 4:** Panel evaluation of any queries scored as potentially harmful by at least one SME. Note that queries only evaluated by a single SME were not included in the fully evaluated 426 queries reporting final scores but were reviewed by the panel out of an abundance of caution.

| #SMEs who scored the query as potentially harmful out of the total # SME evaluators who scored the query | Query Count | Panel agreed with SME potentially harmful score (N) | Panel disagreed with SME potentially harmful score (N) |
| --- | --- | --- | --- |
| 2 of 2 SMEs | 2 | 0 | 2 |
| 1 of 3 SMEs | 25 | 2 | 23 |
| 1 SME (only reviewed by 1 SME) | 8 | 0 | 8 |
| **Total Reviewed by Panel** | **35** | **2** | **33** |

The panel agreed with the ‘No Harm’ rating for 31 of the 35 responses. Of the remaining 4 cases (Supplementary Material 11), the panel *disagreed* with the 2 responses flagged as harmful by both SMEs, determining they were not harmful. However, the panel agreed with a single SME’s assessment on 2 responses that were not flagged by both SMEs, classifying them as potentially harmful. Despite this, the total number of potentially harmful responses remained 2 (0.47%), aligning with the SME-rated results. SME ratings are reported as final; the panel’s role is to validate the reliability of the methodology and to surface edge cases that warrant further product development.

The 33 queries where the panel did not agree with the score for potential harm fell into several categories: 15 responses where the SME expected content beyond the scope of the original query; 10 responses where the SME expressed concern about response clarity where the internal clinical panel process deemed the response appropriate for the intended clinical audience; 6 responses where the SME perceived omissions in the response not perceived as omissions by the internal review process; and 2 responses where the SME preferred the response align with alternate clinical guidelines or interpretations of guidelines, though the internal review panel found the response was aligned with current clinical guidelines.

Among the 426 evaluated query-response pairs, two SMEs agreed on 258 (60.6%). The remaining 168 (39.4%) required a third SME to resolve disagreement, and of those, 17 (10.1%) required email consensus. In 5 of these 17 cases, one SME did not respond during the consensus period, and the lowest score was adopted as the final rating.

Among the five evaluated dimensions, SMEs disagreed on completeness more frequently than other dimensions: 117 (27.5%) query-response pairs required 3^rd^ SME review and represented 12 of the 17 query response-pairs requiring email consensus. Further details on the distribution of agreement, disagreement resolved with a 3rd reviewer, cases requiring email consensus, and final score resolution patterns can be found in Supplementary Material 12 (Tables 1-4).

## DISCUSSION

Deploying GAI products in a clinical setting requires robust evaluation strategies to assess risks to patient care. This case study presents a clinician-in-the-loop methodology for quantifying performance in a real-world application. While the demonstrated evaluation of CK AI provides a guide for assessing a Retrieval Augmented Generation (RAG) system, the five dimension framework applies broadly to other forms of generative AI. Prompt engineered out of the box LLMs, fine tuned models, or systems with text generation capabilities could be assessed on these dimensions with minimal adaptation.

The strengths of this approach are seen across query set development, dimensions and metrics, and SME selection, training and query-response pair assignment. A key advantage of our multi SME, multi dimensional framework is its improved reliability over simpler one or two reviewer models. A multi-evaluator approach using the mode or consensus score was taken to reduce unwarranted misclassification of errors in reporting the performance of the product.

Early in our pilot evaluations, we trialed approaches where an internal clinical panel acted as a single or third reviewer to resolve disagreements, surfacing two key drawbacks: (1) potential bias when internal teams also influenced product development, and (2) insufficient specialty depth compared to external, board certified clinicians. Single evaluator methods can minimize costs and expedite reviews, yet we found this approach risks overlooking nuanced clinical perspectives and underestimates subjective variability in ratings. Although this multi SME approach requires additional time and expense, we reduce the likelihood of one individual’s biases overshadowing final point-estimates.

While this methodology is meant to limit subjectivity, it is critical to avoid overlooking possible concerns about potentially harmful content. To guard against missed safety issues, every query-response pair flagged for harm (n = 35) underwent internal panel review. The panel upheld 2 single-SME flags, overturned the other 31, and disagreed with both SMEs on potential for harmfulness of the query-response pairs agreed upon by 2 SMEs. Accordingly, the harm rate held steady at 2 / 426 (0.47 %). This outcome highlighted the need to further train users and evaluators on the intended use and the reductive limitations of the tool as it summarizes information.

There are important limitations to consider when interpreting results. While this methodology establishes a foundation, several enhancements could address these limitations. First, our primary focus was on the ten most common ABMS specialties, presenting an opportunity for future evaluations targeting rare conditions and niche clinical edge-cases.

Second, the SMEs in this evaluation were compensated contractors of Elsevier, representing a potential conflict of interest. We sought to mitigate this risk by employing multiple independent reviewers, standardized rating guidelines, applying a consensus-based resolution protocol, and blinding evaluators to product metric targets. To mitigate potential biases stemming from SME conflicts of interest, future evaluations could incorporate non affiliated SMEs.

Third, human evaluations of open-ended LLM responses are inherently subjective: each SME may interpret the same query-response pair differently and have unique expectations for the structure and content of the free form responses. Open-ended user queries can be interpreted in various ways both by evaluators and the LLM, leading to subjective assessments of the system. Similarly, evaluators have subjective expectations of the response. The level of SME disagreement underscores this variability. These challenges mirror findings from other “LLM-as-a-judge” studies, where consistency in rating free-form text remains difficult [18]. To reduce subjectivity in scoring open ended responses, more robust SME training and calibrated examples could be employed, along with automated checks. Future evaluations may additionally benefit from developing SME provided ground truth answers for certain query types, particularly where clinically accepted ‘gold standard’ responses exist. Reference answers could help anchor evaluators, mitigate subjective scoring differences, and enable additional quantitative metrics (e.g., ROUGE, BLEU). However, for open ended, context specific queries, we believe complementary human in the loop judgments remain essential for identifying nuanced correctness and harm.

Fourth, the evaluation relied on static query-response pairs, preventing SMEs from asking clarifying follow-up questions or refining their queries, as would typically occur in real-world usage. Interactive testing, where SMEs can submit follow up or clarifying queries would more accurately reflect clinical reality and might reduce perceived omissions. While user experience, including individual preferences and iterative querying, is a vital research area, it was beyond the scope of this study.

Fifth, systematic assessments of demographic bias and citation relevance could complement this framework, particularly if integrated into existing clinician driven evaluation processes. Because biases in LLM responses may disproportionately affect minority populations and can introduce disparities in care, future iterations of this framework should incorporate an additional dimension that explicitly captures demographic and cultural considerations, including potential racial, ethnic, or gender biases.

Finally, a notable limitation is the resource intensity of SME evaluations. On average, one query response pair required approximately 15 minutes for each SME to score. With a total of 426 evaluated queries, with some requiring 3rd SME review, this translates to approximately 255 hours of highly specialized SME time, making this approach less feasible for extremely large datasets or those with cost constraints. Future work should explore semi automated or complementary evaluation methods to reduce these practical burdens. By directly targeting these areas, future evaluations can refine the reliability and scalability of clinical GAI assessments while ensuring their safety and effectiveness in healthcare applications.

This evaluation framework contributes to the body of evidence for assessing GAI in healthcare, while the detailed methodology addresses the critical gap in reproducible implementation guidance. It is an important building block for establishing more efficient approaches to evaluation. As we identify gaps and share findings through frameworks like this one, we move toward comprehensive evaluation best practices that combine the strengths of human clinical judgment with scalable approaches.

## CONCLUSION

In this work, we introduced a clinician in the loop evaluation framework encompassing five core dimensions to systematically assess generative AI outputs. By combining robust human evaluation protocols, standardized rating scales, and structured dispute resolution, we provide a blueprint that integrates objective performance metrics with qualitative clinical expert judgments. Although subjectivity persists and practical constraints remain, this framework can be adapted to offer a scalable path toward more reliable clinical GAI evaluation and safe deployment. Ultimately, by sharing these methods and lessons learned, we aim to advance reproducible best practices for safely integrating generative AI tools into healthcare.

## DATA AVAILABILITY STATEMENT

The source code and data, including all queries, responses, and ratings, underlying this article are available in Dryad (supplementary material 7-10)[31]. While not assessed as Protected Health Information, which may be subject to regulatory protections, some queries contained patient case information. We evaluated the accuracy of the responses to these queries but due to the potential sensitivity of the data we redacted some of these queries and responses from the published data sets.

## CONTRIBUTORSHIP STATEMENT

LL co-developed the evaluation framework, directed the team, oversaw analysis, and wrote sections of the manuscript. AFU co-developed the evaluation framework, oversaw the alignment of the framework to clinical best practices, and wrote sections of the manuscript. AB co-completed the data analysis and co-developed the evaluation framework and wrote sections of the manuscript. KB: co-completed the data analysis and co-developed the evaluation framework. TM oversaw the SME recruitment, education, onboarding, management, and facilitated the SMEs evaluation activities. DH co-developed the evaluation framework and oversaw the technical implementation of the framework VA co-served as the technical owner of the evaluation framework including additional future refinements.

## ACKNOWLEDGEMENTS

We would like to thank Payal Mitra, Louise Chang, Angela Anderson, Katie Scranton, and Mie-Yun Lee who contributed significantly to an early version of the dimensions in the evaluation prototype and executed prototype evaluation rounds that informed the development of this framework. We would also like to thank Ashley Fowler, Harsh Sindhwa, Pranita Mahajan, Radha Rathore, Vijay Somanath, Vidhyaa Sankagiri, and Sameer Chivukula, in addition to the entire evaluation team who facilitated the execution of several rounds of evaluation of multiple products that helped to improve the framework and execution. Lastly, we would like to thank the clinical evaluators who did the important work of assessing responses, and we express our sincere gratitude to the reviewers of this manuscript who provided valued feedback.

## REFERENCES

1. Meng X, Yan X, Zhang K, et al. The application of large language models in medicine: A scoping review. iScience 2024;27(5):109713. doi: 10.1016/j.isci.2024.109713.

2. Vrdoljak J, Boban Z, Vilovik M, et al. A review of large language models in medical education, clinical decision support, and healthcare administration. Healthcare 2025;13,603. DOI: 10.3390/healthcare13060603.

3. Ng KKY, Matsuba I, Zhang PC. RAG in Health Care: A Novel Framework for Improving Communication and Decision-Making by Addressing LLM Limitations. NEJM AI 2025;2(1). DOI: 10.1056/AIra2400380.

4. Alkaissi H, McFarlane SI. Artificial Hallucinations in ChatGPT: Implications in Scientific Writing. Cureus 2023 Feb 19;15(2):e35179. doi: 10.7759/cureus.35179. PMID: 36811129; PMCID: PMC9939079.

5. Li Y, Li J, Li M, et al. VaxBot-HPV: a GPT-based chatbot for answering HPV vaccine-related questions. JAMIA Open 2025;8(1). DOI: 10.1093/jamiaopen/ooaf005.

6. Li Y, Zhao J, Li M, et al. RefAI: a GPT-powered retrieval-augmented generative tool for biomedical literature recommendation and summarization. JAMIA 2024;31(9). DOI: 10.1093/jamia/ocae129

7. Zakka C, Shad R, Chaurasia A, et al. Almanac — Retrieval-Augmented Language Models for Clinical Medicine. NEJM AI 2024;1(2). DOI: 10.1056/aioa2300068.

8. Amugongo L, Smith T, Johnson R, et al. Retrieval Augmented Generation for Large Language Models in Healthcare: A Systematic Review. researchgate.net. [Online] July 2024. [Cited: September 4, 2024.] https://www.researchgate.net/publication/382227452_Retrieval_Augmented_Generation_for_Large_Language_Models_in_Healthcare_A_Systematic_Review.

9. Lee P, Bubeck S, Petro J. Benefits, limits, and risks of GPT-4 as an AI chatbot for medicine. NEJM 2023;338(13). doi: 10.1056/NEJMsr2214184.

10. Lee M. A Mathematical Investigation of Hallucination and Creativity in GPT Models. Mathematics 2023;11(10), 2320. doi: 10.3390/math11102320.

11. Li Y, Li F, Roberts K, et al. A Comparative Study of Recent Large Language Models on Generating Hospital Discharge Summaries for Lung Cancer Patients [preprint]. 2024. Available from: https://arxiv.org/abs/2411.03805 (accessed 8 April 2025)

12. Papineni K, Roukos S, Ward T, et al. Bleu: a method for automatic evaluation of machine translation. In: Proceedings of the 40th Annual Meeting of the Association for Computational Linguistics; 2002 Jul 6–12; Philadelphia, PA. Stroudsburg (PA): Association for Computational Linguistics; 2002. p. 311–318.

13. Chin-Yew Lin. ROUGE: a package for automatic evaluation of summaries. In: Text Summarization Branches Out, 2004; Barcelona, Spain: Association for Computational Linguistics; 2004. p. 74–81.

14. Liang P, Bommasani R, Lee T, et al. Holistic Evaluation of Language Models. Transactions on Machine Learning Research (TMLR) 2023. arXiv:2211.09110 10.48550/arXiv.2211.09110

15. Tam TYC, Sivarajkumar S, Kapoor S, et al. A framework for human evaluation of large language models in healthcare derived from literature. npj Digit Med 2024;7:258. 10.1038/s41746-024-01258-7.

16. Wei Q, Yao Z, Cui Y, et al. Evaluation of ChatGPT-Generated Medical Responses: A Systematic Review and Meta-Analysis. J Biomed Inform 2024 Mar;151:104620. doi: 10.1016/j.jbi.2024.104620.

17. Park YJ, Pillai A, Deng J, et al. Assessing the research landscape and clinical utility of large language models: a scoping review. BMC Med Inform Decis Mak 2024 Mar 12;24(1):72. doi: 10.1186/s12911-024-02459-6. PMID: 38475802; PMCID: PMC10936025.

18. Johri S, Jeong J, Tran BA, et al. An evaluation framework for clinical use of large language models in patient interaction tasks. Nat Med 2025 Jan;31(1):77–86. doi: 10.1038/s41591-024-03328-5. Epub 2025 Jan 2. PMID: 39747685.

19. Singhal K, Tu T, Gottweis J, et al. Towards expert-level medical question answering with large language models [preprint]. 2023. Available from: https://arxiv.org/pdf/2305.09617v1 (accessed 8 Nov 2023).

20. Food and Drug Administration. Artificial intelligence and machine learning (software as a medical device). [Online]. Available at: https://www.fda.gov/medical-devices/software-medical-device-samd/artificial-intelligence-and-machine-learning-software-medical-device (accessed 28 Oct 2024).

21. Assurance Standards Guide and Reporting Checklist. Coalition for Health AI (CHAI). [Online] June 6, 2024. Available from: https://chai.org/assurance-standards-guide/ (accessed 4 September 2024).

22. Article 92: Power to Conduct Evaluations. artificialintelligenceact.eu. [Online]. Available at: https://artificialintelligenceact.eu/article/92/#:~:text=The%20AI%20Office%2C%20after%20consulting,testing%20and%20risk%20prevention%20measures (accessed 17 Oct 2024).

23. Elsevier. [Online] https://www.elsevier.com/about/press-releases/elsevier-health-launches-clinicalkey-ai-the-most-advanced-gen-ai-powered (accessed January 2025).

24. Marshall G, Hays R. The Patient Satisfaction Questionnaire Short Form (PSQ-18). RAND Corporation. Available at: http://www.rand.org/content/dam/rand/pubs/papers/2006/P7865.pdf (accessed 8 November 2024).

25. Larsen DL, Attkisson CC, Hargreaves WA, et al. Assessment of client/patient satisfaction: development of a general scale. Eval Program Plann 1979;2(3):197–207. doi: 10.1016/0149-7189(79)90094-6.

26. Hoppes M, Mitchell JL, Venditti EG, et al. Serious safety events: Getting to Zero™. J Health Risk Manag 2013;32(3):27–45. doi: 10.1002/jhrm.21098. PMID: 23335299.

27. American Board of Medical Specialties. ABMS Board Certification Report, 2022–2023. [Place unknown]: ABMS; 2023. Available at: https://www.abms.org/wp-content/uploads/2023/11/abms-board-certification-report-2022-2023.pdf (accessed 17 Oct 2024).

28. [dataset]* Zakka C, Chaurasia A, Shad R, et al. Almanac: retrieval-augmented language models for clinical medicine [dataset/preprint]. arXiv 2023:2303.01229. Available at: https://arxiv.org/abs/2303.01229 (accessed 17 Oct 2024).

29. Ely JW, Osheroff JA, Gorman PN, et al. A taxonomy of generic clinical questions: classification study. BMJ 2000 Aug 12;321(7258):429–32. doi: 10.1136/bmj.321.7258.429. PMID: 10938054; PMCID: PMC2

30. Stone Fish L, Busby DM. The Delphi method. In: Sprenkle DH, Piercy FP, eds. Research methods in family therapy. 2nd ed. New York: The Guilford Press 2005:238–53. Available at: https://psycnet.apa.org/record/2005-08638-013 (accessed 17 Oct 2024).

