## Supplementary Material 1 for "Reproducible Generative AI Evaluation for Healthcare: A Clinician-in-the-Loop Approach"

**Supplementary Material 1: Guidelines for curating and labeling queries**

The queries were labeled by internal physicians and the list was curated prior to running the queries on CK AI.

All queries should be **relevant at the point of care** for a physician. This requirement is met by pulling and reviewing queries from current users of the product or by using queries approved by internal physicians which are relevant to point of care and the intended use of the product.

Queries should represent the **most common areas of clinical practice** in the US. This requirement was met by internal physicians labeling queries with as many specialties as should have the expertise to answer the query based on the topic. Additional 318 non-randomly chosen queries were added from point of care queries created by internal and external clinical consultants where there were less than 20 per specialty, or to approximate the specialty distribution in any of the top 10 most common specialty areas as outlined by the ABMS.

A random set of 700 user queries entered between May 1 – September 1, 2024, from all markets and all users were obtained. The list was cleaned to exclude queries entered by internal users, non-English queries, duplicates, incomplete thoughts (including, “retained surgical,” “what can I do if my”; “Neut# <1.0”; “can person reinfected in wnv”; “abs”; “what should be done for a patient G”), unrelated to medical care (including “should I slow dance with ibuprofen”; “how will I know when to retire from healthcare”, “use of bioactive materials in dentistry”), non-human related queries (including “should cat with fever of unknown origin get antibiotics”). Keyword searches, labeled with the query type 6.1.1.1, unclassified, defined as entries without a clear question word or a single question that could be assumed from the entry, were removed (including, “Class 2 obesity”, “syphilis diagnosis”, “bat exposure”, “PRP and CMC OA”, “Erythema marginatum”, “pain in front of knee cap”, “Barostim procedure”, etc.).

| [From ABMS Board Certification Report, 2022-2023](https://www.abms.org/wp-content/uploads/2023/11/abms-board-certification-report-2022-2023.pdf) | **Total Active Physicians** | **Percent of physician total** |
| --- | --- | --- |
| **Required Specialties** | **n** | **% of total** |
| Internal Medicine | 262,046 | 26% |
| Pediatrics | 115,131 | 11% |
| Family Medicine/General Practice | 100,794 | 10% |
| Radiology | 65,151 | 6% |
| Anesthesiology | 60,880 | 6% |
| Psychiatry | 59,719 | 6% |
| Obstetrics and Gynecology | 54,545 | 5% |
| General Surgery | 44,758 | 4% |
| Emergency Medicine | 43,927 | 4% |
| Pathology | 33,033 | 3% |

### **Gender** labels were added prior to evaluation, but no specific number was required. Non-binary genders were labelled as part of special populations.

**Ages** were labelled by category, only when the query specified an age (range). No specific number of queries were required per category of age. Categories included: Premature, Neonate, Infant, Child, Adolescent, Adult, and Geriatric.

**Special population** labels were added, only when the query specified. No specific number of queries were required per special population. Categories included:

1. Pregnant and breastfeeding
2. Infertile
3. People with disabilities and/or rare disorders
4. Rural populations
5. Underrepresented ethnic/racial populations (African American, Latinos, Native Americans, etc.)
6. Non-US-born
7. Non-English speaking
8. Populations with low socioeconomic status
9. Food desert/food insecurity
10. LGBTQ+
11. Religious groups
12. Homeless
13. Veterans
14. Day Laborers
15. Migrant Farmworkers
16. Asylum Seekers
17. Sex Workers
18. Victims of trafficking
19. Single Parent

**Sensitive topic** labels were added, only when the query specified. No specific number of queries were required per special population. Categories included:

1. HIV and other STIs
2. Alcohol/substances
3. Obesity
4. Mental Health/Suicidality
5. Medical misinformation (e.g., antivaccine)
6. COVID
7. Abortion
8. Domestic or sexual violence
9. Pain management
10. Palliative care/End of Life

**Query Type** was labeled based on categories presented by John Ely’s paper published in BMJ in 2000 (18) which introduced a query taxonomy, classifying questions in primary care the categories of diagnosis, treatment, management, epidemiology and nonclinical, with the following subcategories:

1.1.1.1 --- diagnosis --- cause/interpretation of clinical finding --- symptom
        1.1.2.1 --- diagnosis --- cause/interpretation of clinical finding --- sign
        1.1.3.1 --- diagnosis --- cause/interpretation of clinical finding --- test finding (lab, ECG, imaging, biopsy, skin test, etc.)
        1.1.4.1 --- diagnosis --- cause/interpretation of clinical finding --- unspecified findings or multiple categories of findings
        1.2.1.1 --- diagnosis --- criteria/manifestations
        1.3.1.1 --- diagnosis --- test (lab, skin test, biopsy, imaging, element of physical exam, etc.) --- indications/efficacy
        1.3.2.1 --- diagnosis --- test (lab, ECG, imaging, biopsy, skin test, element of physical exam, etc.) --- accuracy
        1.3.3.1 --- diagnosis --- test (lab, ECG, imaging, biopsy, skin test, element of physical exam, etc.) --- timing/monitoring
        1.3.4.1 --- diagnosis --- test (lab, ECG, imaging, biopsy, skin test, element of physical exam, etc.) --- preparation
        1.3.5.1 --- diagnosis --- test (lab, ECG, imaging, biopsy, skin test, element of physical exam, etc.) --- method
        1.4.1.1 --- diagnosis --- name finding --- body part (anatomy) on physical exam or imaging study
        1.4.2.1 --- diagnosis --- name finding --- condition
        1.4.3.1 --- diagnosis --- name finding --- test
        1.5.1.1 --- diagnosis --- orientation --- condition
        1.5.2.1 --- diagnosis --- orientation --- test
        1.6.1.1 --- diagnosis --- inconsistencies
        1.7.1.1 --- diagnosis --- cost
        1.8.1.1 --- diagnosis --- not elsewhere classified
        2.1.1.1 --- treatment --- drug prescribing --- how to prescribe --- undifferentiated
        2.1.1.2 --- treatment --- drug prescribing --- how to prescribe --- dosage
        2.1.1.3 --- treatment --- drug prescribing --- how to prescribe --- timing
        2.1.2.1 --- treatment --- drug prescribing --- efficacy/indications/drug of choice --- treatment
        2.1.2.2 --- treatment --- drug prescribing --- efficacy/indications/drug of choice --- prevention
        2.1.3.1 --- treatment --- drug prescribing --- adverse effects --- findings caused by drug/adverse effects of drug
        2.1.3.2 --- treatment --- drug prescribing --- adverse effects --- administration in face of adverse effects
        2.1.3.3 --- treatment --- drug prescribing --- adverse effects --- safety/contraindications (includes pregnancy and breast feeding)
        2.1.4.1 --- treatment --- drug prescribing --- interactions
        2.1.5.1 --- treatment --- drug prescribing --- name finding
        2.1.6.1 --- treatment --- drug prescribing --- orientation/composition
        2.1.7.1 --- treatment --- drug prescribing --- physical characteristics
        2.1.8.1 --- treatment --- drug prescribing --- pharmaco-dynamics/absorption
        2.1.9.1 --- treatment --- drug prescribing --- mechanism of action
        2.1.10.1 --- treatment --- drug prescribing --- cost
        2.1.11.1 --- treatment --- drug prescribing --- serum levels
        2.1.12.1 --- treatment --- drug prescribing --- availability
        2.2.1.1 --- treatment --- not limited to but may include drug prescribing --- efficacy/indications --- treatment
        2.2.1.2 --- treatment --- not limited to but may include drug prescribing --- efficacy/indications --- prevention
        2.2.2.1 --- treatment --- not limited to but may include drug prescribing --- timing
        2.2.3.1 --- treatment --- not limited to but may include drug prescribing --- how to do it
        2.2.4.1 --- treatment --- not limited to but may include drug prescribing --- principles/rationale
        2.3.1.1 --- treatment --- not elsewhere classified
        3.1.1.1 --- management (not specifying diagnostic or therapeutic) --- condition/finding
        3.2.1.1 --- management (not specifying diagnostic or therapeutic) --- other providers --- practices of other providers
        3.2.2.1 --- management (not specifying diagnostic or therapeutic) --- other providers --- referral
        3.2.3.1 --- management (not specifying diagnostic or therapeutic) --- other providers --- community services
        3.3.1.1 --- management (not specifying diagnostic or therapeutic) --- doctor-patient communication --- how to advise
        3.3.2.1 --- management (not specifying diagnostic or therapeutic) --- doctor-patient communication --- how to approach difficult issue
        3.3.3.1 --- management (not specifying diagnostic or therapeutic) --- doctor-patient communication --- patient compliance
        3.4.1.1 --- management (not specifying diagnostic or therapeutic) --- not elsewhere classified
        4.1.1.1 --- epidemiology --- prevalence/incidence
        4.2.1.1 --- epidemiology --- etiology --- causation/association --- risk factors/disease agents
        4.2.1.2 --- epidemiology --- etiology --- causation/association --- genetics
        4.3.1.1 --- epidemiology --- course/prognosis
        4.4.1.1 --- epidemiology --- not elsewhere classified
        5.1.1.1 --- nonclinical --- education --- provider --- continuing medical education
        5.1.1.2 --- nonclinical --- education --- provider --- information source
        5.1.1.3 --- nonclinical --- education --- provider --- trainee
        5.1.2.1 --- nonclinical --- education --- patient
        5.2.1.1 --- nonclinical --- administration
        5.3.1.1 --- nonclinical --- ethics
        5.4.1.1 --- nonclinical --- legal
        5.5.1.1 --- nonclinical --- frustration
        5.6.1.1 --- nonclinical --- not elsewhere classified
        6.1.1.1 --- unclassified
