## Supplementary Material 2 for "Reproducible Generative AI Evaluation for Healthcare: A Clinician-in-the-Loop Approach"

**Supplementary Material 2: Metadata Labels and Distributions**

|  |  | Query Source Set | | Evaluated Queries | |
| --- | --- | --- | --- | --- | --- |
| Dimension | Group | n | % | n | % |
| Age Category | Adolescent | 21 | 3.32 | 21 | 4.93 |
|  | Adult | 60 | 9.48 | 40 | 9.39 |
|  | Child | 28 | 4.42 | 18 | 4.23 |
|  | Geriatric | 14 | 2.21 | 11 | 2.58 |
|  | Infant | 19 | 3.00 | 17 | 3.99 |
|  | Neonate | 10 | 1.58 | 8 | 1.88 |
|  | Premature | 3 | 0.47 | 3 | 0.70 |
|  | Unspecified | 19 | 3.00 | 13 | 3.05 |
|  | N/A | 493 | 77.88 | 328 | 77.00 |
| Primary Specialty | Anesthesia | 38 | 6.00 | 35 | 8.22 |
|  | Emergency Medicine | 35 | 5.53 | 29 | 6.81 |
|  | Family Medicine | 90 | 14.22 | 53 | 12.44 |
|  | General Surgery | 38 | 6.00 | 27 | 6.34 |
|  | Internal Medicine | 207 | 32.70 | 105 | 24.65 |
|  | OB/GYN | 46 | 7.27 | 37 | 8.69 |
|  | Pathology | 20 | 3.16 | 18 | 4.23 |
|  | Pediatrics | 77 | 12.16 | 60 | 14.08 |
|  | Psychiatry | 56 | 8.85 | 51 | 11.97 |
|  | Radiology | 26 | 4.11 | 11 | 2.58 |
| Query Type | Diagnosis | 226 | 35.70 | 148 | 34.74 |
|  | Epidemiology | 61 | 9.64 | 39 | 9.15 |
|  | Management (not specifying diagnostic or therapeutic) | 35 | 5.53 | 23 | 5.40 |
|  | Nonclinical | 17 | 2.69 | 14 | 3.29 |
|  | Treatment | 294 | 46.45 | 202 | 47.42 |
| Sensitive Topics* | Alcohol/substances | 10 | 1.58 | 10 | 2.35 |
|  | COVID | 4 | 0.63 | 1 | 0.23 |
|  | Cancer | 31 | 4.90 | 25 | 5.87 |
|  | Domestic or sexual violence | 2 | 0.32 | 1 | 0.23 |
|  | HIV and other STIs | 2 | 0.32 | 1 | 0.23 |
|  | Mental Health/Suicidality/Eating Disorder | 23 | 3.63 | 22 | 5.16 |
|  | Obesity | 5 | 0.79 | 3 | 0.70 |
|  | Pain management | 5 | 0.79 | 2 | 0.47 |
|  | Palliative care / end of life | 14 | 2.21 | 9 | 2.11 |
|  | N/A | 540 | 85.31 | 354 | 83.10 |
| Sex at Birth | Female | 57 | 9.00 | 43 | 10.09 |
|  | Male | 30 | 4.74 | 15 | 3.52 |
|  | N/A | 546 | 86.26 | 368 | 86.38 |
| Special Population* | Day Laborers | 1 | 0.16 | 0 | 0.00 |
|  | Homeless | 1 | 0.16 | 0 | 0.00 |
|  | Infertile | 3 | 0.47 | 2 | 0.47 |
|  | Migrant Farmworkers | 2 | 0.32 | 0 | 0.00 |
|  | People with disabilities and/or rare disorders | 8 | 1.26 | 7 | 1.64 |
|  | Populations with low socioeconomic status | 1 | 0.16 | 0 | 0.00 |
|  | Pregnant and breastfeeding | 21 | 3.32 | 18 | 4.23 |
|  | Underrepresented ethnic/racial populations (African American, Latinos, Native Americans, etc.) | 1 | 0.16 | 0 | 0.00 |
|  | Veterans | 1 | 0.16 | 0 | 0.00 |
|  | non-US-born | 2 | 0.32 | 0 | 0.00 |
|  | N/A | 595 | 94.00 | 399 | 93.66 |

*Queries may reference multiple subgroups
