## Supplementary Material 4 for "Reproducible Generative AI Evaluation for Healthcare: A Clinician-in-the-Loop Approach"

**Supplementary Material 4: Number of query-response pairs reviewed by each individual SME and for MD’s, their associated board certification(s). For PharmD’s, their stated specialty area(s).**

| **Credential** | **The MD SME’s specialty board-certification(s), or the specialty area of the PharmD** | **Query-response pairs Reviewed (n)** |
| --- | --- | --- |
| MD | Internal Medicine, Pediatrics | 162 |
|  | Colon and Rectal Surgery, General Surgery | 2 |
|  | ENT, General Surgery | 8 |
|  | Internal Medicine | 107 |
|  | Internal Medicine | 158 |
|  | Dermatology | 13 |
|  | Neurology, Neurosurgery | 24 |
|  | Pediatrics | 83 |
|  | Emergency Medicine | 33 |
|  | Anesthesia | 35 |
|  | Neurology, Psychiatry | 53 |
|  | Emergency Medicine | 36 |
|  | Infectious Disease, Internal Medicine | 44 |
|  | Ophthalmology | 2 |
|  | Gastroenterology, Internal Medicine | 39 |
|  | Cardiology | 45 |
|  | Pediatrics | 72 |
|  | Heme/Onc, Internal Medicine | 45 |
|  | Neurology | 25 |
|  | Colon and Rectal Surgery, General Surgery, Thoracic Surgery | 19 |
|  | Dermatology | 14 |
|  | OB/GYN, Preventive Medicine | 45 |
|  | Cardiology, Internal Medicine | 19 |
|  | Ophthalmology, Plastic Surgery | 2 |
|  | Neurology | 26 |
|  | Ophthalmology | 2 |
|  | Allergy and Immunology, Internal Medicine | 22 |
|  | Anesthesia | 36 |
|  | Pathology | 19 |
|  | Pathology | 19 |
|  | ENT | 7 |
|  | Psychiatry | 55 |
|  | OB/GYN | 47 |
|  | Nuclear Medicine, Radiology | 30 |
|  | Psychiatry | 51 |
|  | General Surgery | 13 |
|  | Family Medicine | 69 |
| PharmD | Gastroenterology | 35 |
|  | Dermatology | 13 |
|  | Family Medicine | 7 |
|  | Geriatrics, OB/GYN | 8 |
