## Supplementary Material 5 for "Reproducible Generative AI Evaluation for Healthcare: A Clinician-in-the-Loop Approach"

**Supplementary Material 5: Queries that did not generate a response by CK AI**

**Table 1: automated non-response generating queries producing a user-error (n=4) message for out-of-scope queries, or a ‘no sources’ (n=15) message when relevant content could not be found to produce a response.**

| **Query** | **Status** |
| --- | --- |
| Is clindamycin a macrolide? Does it interact with flecanaide or is it contraindicated for use with flecanaide? | **no_sources** |
| calculate the dosage of tylenol for an 8 year old | **user_error** |
| what is a flutter valve used for | **no_sources** |
| Tell me about the psychological phenomenon of omorashi and urolagnia. Is it connected to gender dysphoria, sexual abuse, or similar? What about the psychological phenomenon of age regression? | **no_sources** |
| does factor 2 impact mentration after blood thinner | **no_sources** |
| is Pluralibacter gergoviae at high risk of inducible resistance and what antibiotics should work against it? | **no_sources** |
| what is the prevalence of OSA in Hong Kong | **no_sources** |
| true sinusoidal efm patter have what characteristics | **no_sources** |
| daily maintenance fluid of a 17 kilogram child | **user_error** |
| how to i calculate the hestia score? | **user_error** |
| Is there any difference between Polygala tenuifolia and Polygala senega? | **no_sources** |
| What are the risks of taking nsaids with a patient with et | **no_sources** |
| DIM cause vaginal bleeding | **no_sources** |
| Ddx for formication | **no_sources** |
| lactic acidosis in black widow bite causes | **no_sources** |
| Differences between Ascaris Lumbricoides and Enterobius Vermicularis? | **no_sources** |
| What is adrenal fatigue? | **no_sources** |
| What is the grading system for Hemangioblastomas? | **no_sources** |
| My patient, who was taking 120 mg of oral Morphine at home, is admitted to the hospital with acute renal failure. How do I convert her Morphine into Hydromorphone? | **user_error** |
