## Supplementary Material 6 for "Reproducible Generative AI Evaluation for Healthcare: A Clinician-in-the-Loop Approach"

**Supplementary Material 6: Aggregated Individual SME ratings for each dimension and rating scale.**

**Table 1: All individual SME ratings; not including email consensus values. Note that queries are counted multiple times in this table – once for each SME that rated the query, even if SMEs agreed by providing the same rating for a single query.**

| **Dimension** | **Rating Scale** | **N Rated by a SME** | **%** |
| --- | --- | --- | --- |
| **Overall Answer Helpfulness** | 0 – unhelpful | 18 | 1.76% |
|  | 1 – just ok | 104 | 10.20% |
|  | 2 – helpful | 898 | 88.04% |
| **Comprehension** | 0 – not comprehended | 8 | 0.78% |
|  | 1 – partially comprehended | 29 | 2.84% |
|  | 2 – completely comprehended | 983 | 96.37% |
| **Correctness** | 0 – completely incorrect | 0 | 0% |
|  | 1 – mostly incorrect | 3 | 0.29% |
|  | 2 – equally correct and incorrect | 21 | 2.06% |
|  | 3 – mostly correct | 81 | 7.95% |
|  | 4 – completely correct | 906 | 88.91% |
|  | not applicable | 8 | 0.79% |
| **Completeness** | 0 – incomplete | 33 | 3.24% |
|  | 1 – adequate | 140 | 13.74% |
|  | 2 – comprehensive | 838 | 82.24% |
|  | not applicable | 8 | 0.79% |
| **Potential Clinical Harm** | 0 – no harm | 991 | 97.16% |
|  | 1 – potential harm | 29 | 2.84% |
| **Potential Harmful Level** | 0 – death | 0 | 0% |
|  | 1 – severe harm | 8 | 0.78% |
|  | 2 – moderate harm | 9 | 0.88% |
|  | 3 – mild harm | 8 | 0.78% |
|  | 4 – no harm (near miss) | 4 | 0.39% |
