## Supplementary Material 12 for "Reproducible Generative AI Evaluation for Healthcare: A Clinician-in-the-Loop Approach"

**Supplementary Material 12: Agreement and Disagreement Descriptive Statistics**

**Table 1: Overview of Evaluation Dimensions and their Agreement/Disagreement Patterns**

This table provides a high-level summary of the evaluation dimensions, showing the overall rates of agreement, disagreement, and email consensus for the evaluated items (n=426).

| **Dimension** | **Initial 2-SME Agreement N (%)** | **Disagreement Requiring 3^rd^ SME Review N (%)** | **Disagreement Resolved with 3rd Reviewer (%)** | **Disagreement Requiring**  **Email Consensus N (%)** |
| --- | --- | --- | --- | --- |
| **Overall Answer Helpfulness** | **351 (82.4%)** | **75 (17.6%)** | 70 (16.4%) | 5 (1.2%) |
| **Comprehension** | **403 (94.6%)** | **23 (5.4%)** | 23 (5.4%) | 0 (0%) |
| **Correctness** | **346 (81.2%)** | **80 (18.8%)** | 72 (16.9%) | 8 (1.9%) |
| **Completeness** | **309 (72.5%)** | **117 (27.5%)** | 105 (24.6%) | 12 (2.8%) |
| **Potential Clinical Harmfulness** | **402 (94.4%)** | **24 (5.6%)** | 24 (5.6%) | 0 (0%) |
| **Potential Harm Level** | **401 (94.1%)** | **25 (5.9%)** | 24 (5.6%) | 1 (0.2%) |

**Table 2: Agreement (with Distribution of Final Results)**

This table shows the cases by dimension where the initial two SMEs agreed on all dimensions, including the breakdown of the final results for each evaluation dimension for these cases.

| **Dimension** | **2-SME**  **Agreement**  **N (% of evaluated query-response pairs)** | **Final Results Distribution**  **N (% of cases with 2-SME agreement for the applicable dimension)** |
| --- | --- | --- |
| **Overall Answer Helpfulness** | 351 (82.4%) | (0) unhelpful – 2 (0.5%)  (1) just ok – 9 (2.6%)  (2) helpful – 340 (96.9%) |
| **Comprehension** | 403 (94.6%) | (0) not comprehended – 2 (0.5%)  (1) partially comprehended – 3 (0.7%)  (2) completely comprehended – 398 (98.8%) |
| **Correctness** | 346 (81.2%) | (0) completely incorrect – 0 (0%)  (1) mostly incorrect – 0 (0%)  (2) equally correct/incorrect – 0 (0%)  (3) mostly correct – 4 (1.1%)  (4) completely correct – 340 (98.3%)  (NA) rated as not comprehended – 2 (0.6%) |
| **Completeness** | 309 (72.5%) | (0) incomplete – 2 (0.65%)  (1) adequate – 9 (2.9%)  (2) comprehensive – 296 (95.8%)  (NA) rated as not comprehended – 2 (0.65%) |
| **Potential Clinical Harmfulness** | 402 (94.4%) | (0) no harm – 401 (99.8%)  (1) potential harm – 1 (0.2%) |
| **Potential Harm Level** | 401 (94.1%) | (NA) no harm – 401 (100%)  (0) death – 0 (%)  (1) severe Harm – 0 (%)  (2) moderate Harm – 0 (%)  (3) mild Harm – 0 (%)  (4) no Harm – 0 (%) |

**Table 3: Disagreement Resolved with a 3rd Reviewer via Mode (with Distribution of Final Results)**

This table presents the cases by dimension where disagreement between two SMEs required a third reviewer to resolve the scores, including the final result distribution for each dimension for these cases.

| **Dimension** | **Disagreement Resolved with 3rd Reviewer via the Mode**  **N (% of evaluated query-response pairs)** | **Final Results Distribution**  **N (% of cases with 3-SME mode agreement for the applicable dimension)** |
| --- | --- | --- |
| **Overall Answer Helpfulness** | 70 (16.4%) | (0) unhelpful – 0 (0%)  (1) just ok – 8 (11.4%)  (2) helpful – 62 (88.6%) |
| **Comprehension** | 23 (5.4%) | (0) not comprehended – 0 (0%)  (1) partially comprehended – 1 (4.3%)  (2) completely comprehended – 22 (95.7%) |
| **Correctness** | 72 (16.9%) | (0) completely incorrect – 0 (0%)  (1) mostly incorrect – 0 (0%)  (2) equally correct/incorrect – 1 (1.4%)  (3) mostly correct – 4 (5.6%)  (4) completely correct – 67 (93%) |
| **Completeness** | 105 (24.6%) | (0) incomplete – 1 (1.0%)  (1) adequate – 14 (13.3%)  (2) comprehensive – 90 (85.7%) |
| **Potential Clinical Harmfulness** | 24 (5.6%) | (0) no harm – 23 (95.8%)  (1) potential harm – 1 (0.2%) |
| **Potential Harm Level** | 24 (5.6%) | (NA) no harm – 23 (95.8%)  (0) death – 0 (0%)  (1) severe Harm – 0 (0%)  (2) moderate Harm – 1 (0.2%)  (3) mild Harm – 0 (0%)  (4) no Harm – 0 (%) |

**Table 4: Disagreement Requiring Email Consensus (with Distribution of Final Results)**

This table displays the 3-way disagreements that required email consensus for resolution, with the final scores broken down by dimension.

| **Dimension** | **Disagreement Requiring Email Consensus (%)** | **Final Results Distribution** |
| --- | --- | --- |
| **Overall Answer Helpfulness** | 5 (1.2%) | (0) unhelpful – 2 (40%)  (1) just ok – 3 (60%)  (2) helpful – 0 (0%) |
| **Comprehension** | 0 (0%) | NA |
| **Correctness** | 8 (1.9%) | (0) completely incorrect – 0 (0%)  (1) mostly incorrect – 1 (12.5%)  (2) equally correct/incorrect – 7 (87.5%)  (3) mostly correct – 0 (0%)  (4) completely correct – 0 (0%)  (NA) rated as not comprehended – 0 (0%) |
| **Completeness** | 12 (2.8%) | (0) incomplete – 7 (58.3%)  (1) adequate – 4 (33.3%)  (2) comprehensive – 1 (8.3%)  (NA) rated as not comprehended – 0 (0%) |
| **Potential Clinical Harmfulness** | 0 (0%) | NA |
| **Potential Harm Level** | 1 (0.2%) | (0) death – 0 (0%)  (1) severe Harm – 1 (100%)  (2) moderate Harm – 0 (0%)  (3) mild Harm – 0 (0%)  (4) no Harm – 0 (0%) |
